# Early onset non-syndromic retinal degeneration due to variants in *INPP5E:* phenotypic expansion of the ciliary gene previously associated with Joubert syndrome

**DOI:** 10.1101/2020.08.24.20179085

**Authors:** Riccardo Sangermano, Iris Deitch, Virginie G. Peter, Rola Ba-Abbad, Emily M. Place, Naomi E. Wagner, Anne B. Fulton, Luisa Coutinho-Santos, Boris Rosin, Vincent Dunet, Ala’a AlTalbishi, Eyal Banin, Ana Berta Sousa, Mariana Neves, Anna Larson, Mathieu Quinodoz, Michel Michaelides, Tamar Ben-Yosef, Eric A. Pierce, Carlo Rivolta, Andrew R. Webster, Gavin Arno, Dror Sharon, Rachel M. Huckfeldt, Kinga M. Bujakowska

## Abstract

**Purpose:** Pathogenic variants in *INPP5E* cause Joubert syndrome, a systemic disorder that can manifest with retinal degeneration among other clinical features. We aimed to evaluate the role of *INPP5E* variants in non-syndromic inherited retinal degenerations (IRDs) of varying severity.

**Methods:** Targeted or genome sequencing were performed in 12 unrelated non-syndromic IRD families from multiple research hospitals. Detailed clinical examination was conducted in all probands. The impact of new likely pathogenic variants was modeled on a tertiary INPP5E protein structure and all the new and published variants were analyzed for their deleteriousness and phenotypic correlation.

**Results:** Fourteen *INPP5E* rare alleles were detected, 12 of which were novel. Retinal degeneration in all 12 probands was clinically distinguishable on the basis of onset and severity into Leber congenital amaurosis (n=4) and a milder, later-onset rod-cone dystrophy (n=8). Two probands showed mild ciliopathy features that resolved in childhood. Analysis of the combined impact of both alleles in syndromic and non-syndromic *INPP5E* patients did not reveal clear genotype-phenotype correlation, suggesting involvement of genetic modifiers.

**Conclusions:** The study expands the phenotypic spectrum of disorders due to pathogenic variants in *INPP5E* and describes a new disease association with previously underdiagnosed forms of early-onset non-syndromic IRD.

## INTRODUCTION

Inherited retinal degenerations (IRDs) are a group of genetically heterogeneous disorders characterized by progressive photoreceptor loss due to genetic defects in ^~^270 genes.^1^ Clinically, IRDs may manifest either as an isolated phenotype (non-syndromic IRD) or as a clinical feature of a syndrome, such as ciliopathies that involve multiple organs and tissues, including central nervous system, skeletal and reproductive system, kidney, liver, pancreas, lung, and neuroretina.^2^

Pathogenic variants leading to ciliopathies occur in genes playing either a structural or a functional role in the primary cilium, a specialized organelle protruding from most post-mitotic cells. Cilia act as antennae that “sense” the physical and biochemical stimuli of the cellular environment to promptly initiate the signaling cascades in response of those changes.^3^ Primary cilia play an important role during embryogenesis and organ development and, therefore, ciliary dysfunction can often leads to congenital or early onset disease.^2^ The photoreceptor outer segment is regarded as a specialized primary cilium detecting light stimuli and thus multiorgan ciliopathies often involve retina.^4^ Joubert syndrome-related disorders (JSRD) are an example of ciliopathy with retinal involvement. JSRD are genetically heterogeneous autosomal or X-linked recessive disorders, comprised of abnormal development of the mid-hindbrain (the core Joubert syndrome (JBTS, OMIM#213300)) and one or multiple extra-neurological findings such as retinal degeneration, coloboma, skeletal abnormalities, cystic kidney disease, liver fibrosis, endocrinological disorders.^5^ The diagnostic hallmark of JBTS is the “molar tooth sign,” a radiological finding detectable on axial magnetic resonance imaging of the brain.^6^ Other neurological features include hypotonia/ataxia and developmental delay, irregular breathing patterns, abnormal eye movements, oculomotor apraxia, and intellectual disability.

Pathogenic variants in the *Inositol Polyphosphate-5-Phosphatase E* gene *(INPP5E)* on chromosome 9 are a known cause of JBTS. *INPP5E* is a widely expressed ciliary gene,^7^ encoding a 72-kDa (644 amino acid) phosphatase that plays a critical role in controlling ciliary growth and stability via the phosphoinositide 3-kinase signaling pathway.^8^ INPP5E selectively cleaves the 5^th^ position phosphate from phosphatidylinositol 3,4,5-trisphosphate (PIP3) and phosphatidylinositol 4,5-bisphosphate (PIP2).^9,10^ To date, 26 pathogenic *INPP5E* alleles have been reported in patients with syndromic IRD associated with JBTS, JSRD, or MORM (Mental retardation, truncal obesity, retinal dystrophy and micropenis) syndrome (OMIM#610156).^8,11,12^ Here, we report 18 mostly non-syndromic IRD patients from 12 unrelated families with pathogenic variants in *INPP5E*, thus expanding the phenotypic spectrum of *INPP5E*-associated disease.

## METHODS

### Ethics statement

The study was approved by the institutional review board of all participating institutions (Partners Healthcare System for families OGI2307 3818, OGI1819_3159, OGI2386_3945, the Boston Children’s Hospital Committee on Clinical Investigation for family OGI3559_5164, Instituto de Oftalmologia Dr. Gama Pinto for families LL135, LL105, LL235, the Institutional Review Boards and ethics committees of Moorfields Eye Hospital for families GC19652, GC16358, GC22740, the institutional review board at Hadassah-Hebrew University Medical Center for family MOL0641-1, the Ethics Committee at Rambam Health Care Campus for family TB315_R693) and adhered to the Declaration of Helsinki. Informed consent was obtained from all individuals on whom genetic testing and further molecular evaluations were performed.

### Clinical evaluation

Twelve probands with autosomal recessive retinal degeneration were genotyped. Four were ascertained from two different medical centers in Boston, USA (Massachusetts Eye and Ear and Boston Children’s Hospital), three in the United Kingdom (Moorfields Eye Hospital), three in Portugal (Instituto de Oftalmologia Dr. Gama Pinto), and two in Israel (Hadassah-Hebrew University Medical Center, Rambam Health Care Campus).

Clinical evaluation was performed by experienced ophthalmologists according to previously published protocols and included functional and structural assessments.^13–16^

For proband LL135, brain MRI was performed using a GE Signa HDxt 1.5 T scanner (GE Medical Systems, Milwaukee, WI). The JBTS and control cases were scanned on a 3 T scanner (Verio and Vida, Siemens Healthcare, Erlangen, Germany). Scanning protocols included unenhanced 3D T1 weighted Imaging and T2 spin echo weighted imaging, which were sufficient to make a first diagnosis.

### Genetic analysis

Blood samples were obtained from probands, and when possible their parents, affected, and unaffected siblings. DNA was isolated from peripheral blood lymphocytes by standard procedures. Four probands (OGI2307_3818, OGI1819_3159, OGI2386_3945, and OGI3559_5164) were sequenced using the Genetic Eye Disease (GEDi) panel, described previously.^17^ The GEDi version used in this study (v6) targeted exons of 278 known IRD genes (Table S1).^1^ The NGS data from the GEDi panel was analyzed using Genome Analysis Toolkit (GATK) version 3 ^18^ and annotated using the Variant Effect Predictor (VEP) tool^19^ with additional annotations taken from the Genome Aggregation Database (GnomAD), the Genomic Evolutionary Rate Profiling (GERP), SIFT, PolyPhen2, CADD and retinal expression.^20–25^ Exome sequencing (ES) for five probands was performed at different facilities (MOL0641-1, Pronto Diagnostics Ltd; LL105, LL135 and LL235, Novogene (HK); TB315_R693, Otogenetics Corporation), as previously described.^15,26,27^ Finally, three patients (GC19652, GC16358, GC22740) underwent genome sequencing (GS, Genomics England) according to previously published protocols.^27^

### Variant validation and phasing

All presented variants refer to the *INPP5E* transcript NM_019892.5. Variant segregation was performed by Sanger sequencing (primers in Table S2) or analysis of NGS reads. For OGI1819_3159, the three *INPP5E* variants detected were phased by cloning and Sanger sequencing (Fig. S1). Briefly, genomic DNA from the proband was amplified using Takara-LA (Takara Bio USA, Inc.) and primers spanning the region containing all variants. The amplified fragment was then cloned into the pCR2.1 plasmid, TA cloning kit (Invitrogen) and Sanger sequenced. Sanger sequencing was performed on ABI 3730xl (Applied Biosystems) using BigDye Terminator v3.1 kits (Life Technologies). Sequence analysis was done using SeqManPro (Lasergene, DNAStar Madison, WI, USA), in which variants were considered to be *in trans* when they were never present on the same clone.

### Multiple sequence alignment, protein modelling, and prediction of missense variants

Multiple sequence alignment of the human INPP5E protein and 99 orthologues was generated using Clustal Omega (https://www.ebi.ac.uk/Tools/msa/clustalo/) and sequences were retrieved from the UniProt Knowledgebase (UniProtKB, https://www.uniprot.org/help/uniprotkb). Tridimensional structure of the INPP5E protein, its putative catalytic sites, and mutated residues were generated with a protein modelling software (PyMOL Molecular Graphics System, Version 1.2r3pre, Schrödinger, LLC) using crystal structure of human INPP5E as an input (Protein Data Bank (PDB) ID: 2XSW). The impact of missense variants on INPP5E structure and function, was predicted by using four prediction algorithms: SIFT (https://sift.bii.a-star.edu.sg/), PolyPhen-2 (http://genetics.bwh.harvard.edu/pph2/), Missense3D (http://www.sbg.bio.ic.ac.uk/~missense3d/) and SuSPect (http://www.sbg.bio.ic.ac.uk/suspect. NetPhos 3.1 Server (http://www.cbs.dtu.dk/services/NetPhos/) was used to predict phosphorylation sites.

## RESULTS

### Novel *INPP5E* variants associated with non-syndromic early onset IRD

Sequencing analysis of 12 recessive non-syndromic IRD families revealed 14 likely pathogenic alleles in *INPP5E* (Fig. 1, Table S3). Twelve alleles were novel, including one complex allele p.[(Ser249Phe);(Arg596Thr)]. All *INPP5E* variants were biallelic, rare (AF≤0.0001 in gnomAD), had high CADD scores (>20) and were predicted to be disease-causing by several *in silico* prediction algorithms (Table S3 and S4). No other variants in IRD genes segregating with the phenotype were found in the probands reported here.

**Fig. 1.**
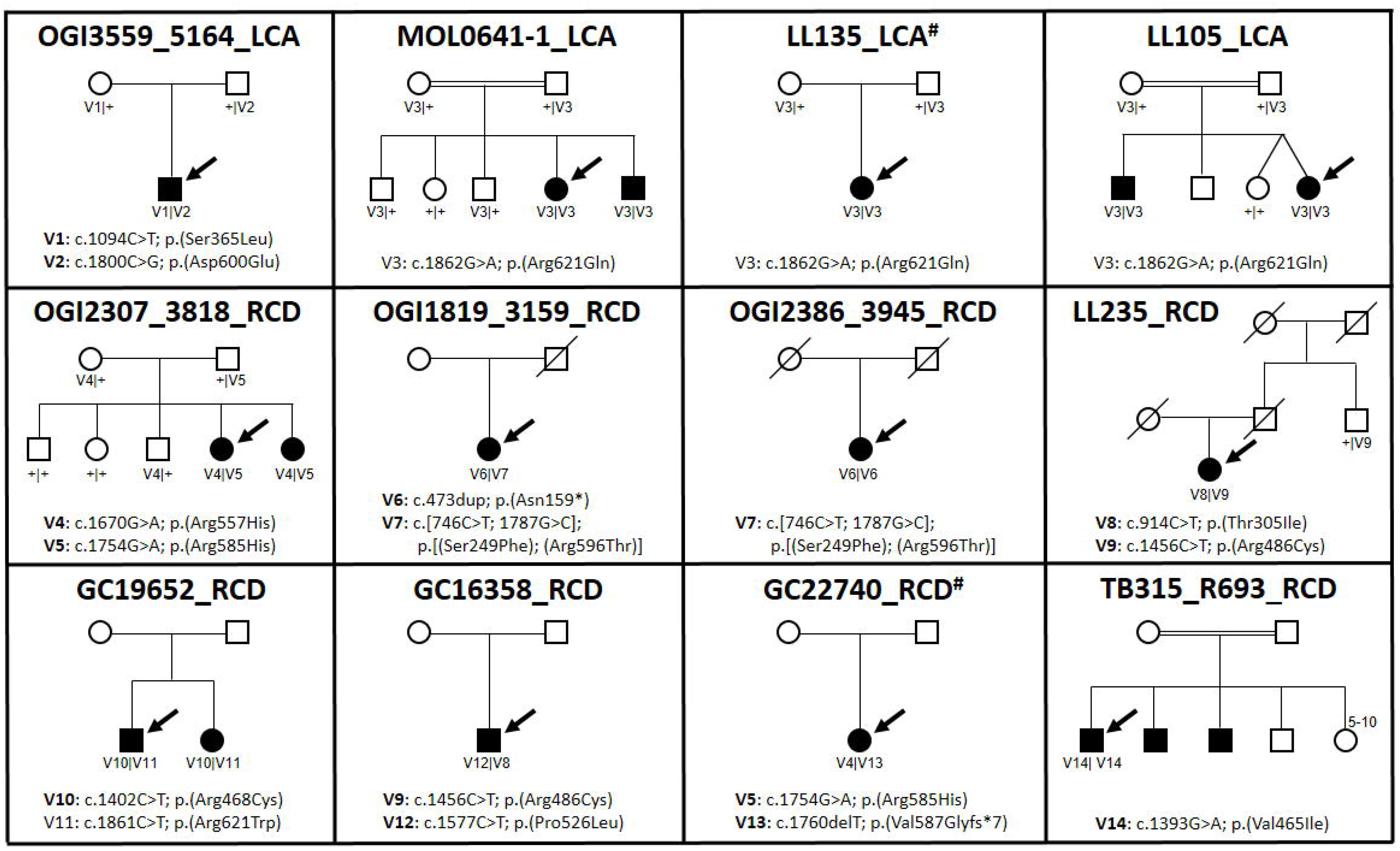
Pedigrees of the 12 non-syndromic IRD families harboring likely pathogenic *INPP5E* variants. Each family is named after the ID of its proband and the specific IRD phenotype diagnosed. Affected male and female subjects are represented with black squares or circles, respectively. Probands are indicated by a black arrow. Novel variants are indicated in bold. When performed, segregation of the *INPP5E* variants in other family members is shown. First cousin marriage is indicated by a double-line. All presented variants refer to the *INPP5E* transcript NM_019892.5.

Among the identified variants, two were protein-truncating: (p.(Asnl59*) and p.(Val587Glyfs*7)), while the remaining were missense. Most of the identified variants clustered in the highly conserved C-terminal half of the protein, containing the inositol 5-phosphatase catalytic domain (residues 273-621) with only two variants (p.(Asnl59*) and p.(Ser249Phe)) located at the N-terminal half of the protein (Fig. 2A, Fig. S2). Consistent with other studies, missense changes mainly affected arginine residues (Fig. 2).^11,29^

**Fig. 2.**
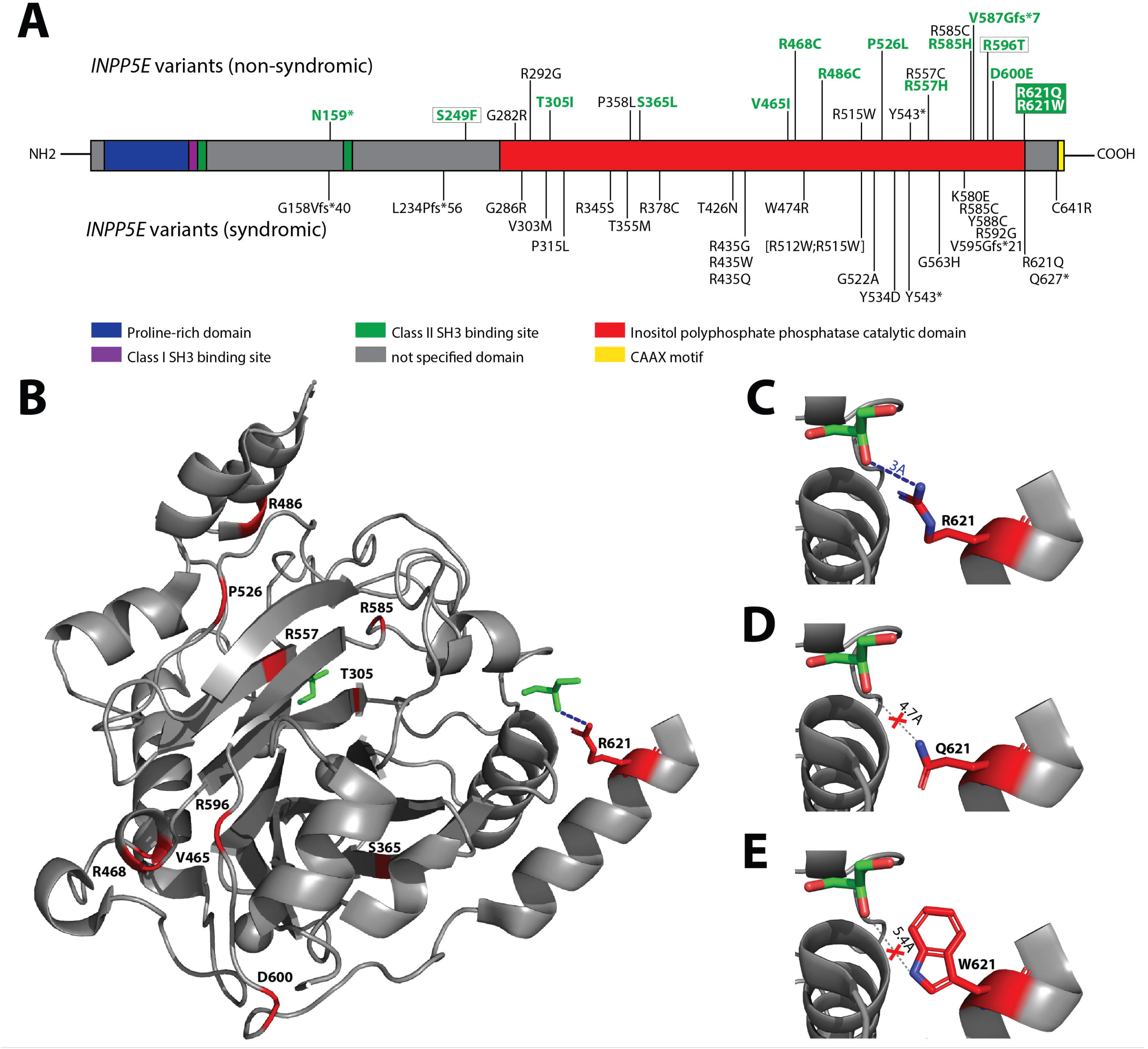
INPP5E structure and protein variants. **(A)** INPP5E secondary structure and distribution of known causal variants. Multiple sequence alignment-derived INPP5E motifs and catalytic domain were highlighted using different colors, while variants were divided in two groups, depending whether they were found in syndromic or non-syndromic IRD patients. The novel variants described in this study are highlighted in bold green, while variants p.(Arg621Gln) and p.(Arg621Trp), found in both our patients and syndromic cases, are indicated by green boxes. Variants p.(Ser249Phe) and p.(Arg596Thr), found to be part of the same complex allele, are indicated by white boxes. **(B)** INPP5E tertiary structure. Tridimensional structure was predicted only for C-terminal 349 amino acids (residues 275-623) available on PDB, as the N-terminal half was classified as disordered region. Two glycerol molecules, acting as proxy for the larger ligand of this protein (i.e., phosphatidylinositol polyphosphate), are shown in green. Amino acid residues for which missense variants in our patients were found are highlighted in red, except for Serine 249 located in the un-modelled region. **(C-E)** Predicted effect of missense variants p.(Arg621Gln) and p.(Arg621Trp) on ligand binding. In the wild-type protein model, Arginine 621 is located in close proximity (3Å) to one glycerol molecule, with which uniquely interacts by establishing one ion bond, indicated by a blue dashed line (C). Missense variants introducing Glutamine (D) or Tryptophan (E) are predicted to increase distance with the glycerol of 4.7Å and 5.4Å, respectively, thus disrupting the ion bond. All presented variants refer to the *INPP5E* transcript NM_019892.5.

Five families (MOLC1641-1, LL105, LL135, OGI2386-3945, TB315_R693) carried homozygous variants (p.(Arg621Gln), p.[(Ser249Phe); (Arg596Thr)], p.(Val465Ile)), and in three of them the parents were first cousins. Five affected members of three unrelated Leber congenital amaurosis (LCA) families with different ethnicities were homozygous for p.(Arg621Gln), thus making this allele the most recurrent *INPP5E* variant found in our study group. Other recurrent missense variants were p.(Arg585His) in OGI2307_3818 and GC22740, p.(Arg486Cys) in LL235, GC19652 and GC16358, and p.[(Ser249Phe);(Arg596Thr)] in OGI1819_3159 and OGI2386_3945.

Variants c.746C>T; p.(Ser249Phe) and c.1787G>C; p.(Arg596Thr) belonged to the same complex allele and were identified in two unrelated patients, homozygous in OGI2386_3945 and compound heterozygous in OGI1819_3159. Although the allele frequency of p.(Ser249Phe) was 11-times higher (AF=0.000067) than of p.(Arg596Thr) (AF=0.000006) (Table S3), based on frequency alone it is not possible to determine which of the variants or both contribute to disease. Ser249 was predicted to be a phosphorylation site for the Protein Kinase C (NetPhos score=0.84, intervals 0-1, Table S4), whereas Arg596 lies in the catalytic domain, though no specific effect of the p.(Arg596Thr) change was predicted.

Alignment of INPP5E protein sequence in 100 species revealed that ten missense changes were affecting highly conserved amino acids (identical in ≥ 98 of species). Three missense variants (p.(Ser249Phe), p.(Ser365Leu), p.(Pro526Leu)) affected less conserved residues that were identical in 84, 70, and 46 species, respectively (Fig. S2).

### Protein modelling and prediction of missense variants at catalytic sites

Modelling of the tertiary structure of INPP5E predicted two sites of potential interaction with a ligand (glycerol molecule used as a proxy of inositol-3-phosphate) (Fig. 2B). The first interaction site resides in the known catalytic domain where the ligand is predicted to form polar bonds with residues His424, Asn479, Asp477, and His584 (Fig. S3). Three of the likely pathogenic variants identified in this study are located either within the catalytic pocket: p.(Arg557His) and p.(Arg585His) or in its proximity: p.(Thr305Ile) (Fig. 2B, Fig. S3). They have the highest score for deleteriousness according to SuSPect^30^ (Table S4). The p.(Thr305Ile) change leads to the disruption of the hydrogen bond connecting Gln339 and Thr305 residues, which is thought to result in the alteration of the INPP5E structure (Fig. S4). None of these new or published likely pathogenic *INPP5E* alleles directly affected the residues predicted to bind the IP3 ligand. The second potential ligand interaction site resides outside of the known catalytic domain and exclusively involves the Arg621 residue. Two of the *INPP5E* variants detected in our patients (p.(Arg621Gln) and p.(Arg621Trp)) targeted the Arg621 residue. Modelling of the structural changes induced by these two variants showed that both lead to disruption of the polar bond connecting the second glycerol molecules to INPP5E protein (Fig. 2C-E).

### Clinical phenotypes

Eight females and four males with *INPP5E*-associated disease demonstrated features of IRDs that could be separated into two clinical categories. Four individuals (OGI3559_5164, M0L0641-1, LL105, LL135) had a more severe retinal degeneration manifested during early infancy (LCA) whereas the remaining eight had a milder juvenile-onset rod-cone degeneration (RCD) (Table 1). All individuals with LCA had nystagmus as a shared early feature. All four subjects had reduced visual acuity with severely constricted visual fields and undetectable or severely reduced elctroretinograms (ERGs). Fundus examination and imaging showed macular and peripheral retinal atrophy. At least two LCA patients (M0L0641-1 and LL135) had structure-function dissociation based on better foveal structure on optical coherence tomography (OCT) than would be expected from their visual acuities (Fig. 3, Table 1).

**Fig. 3.**
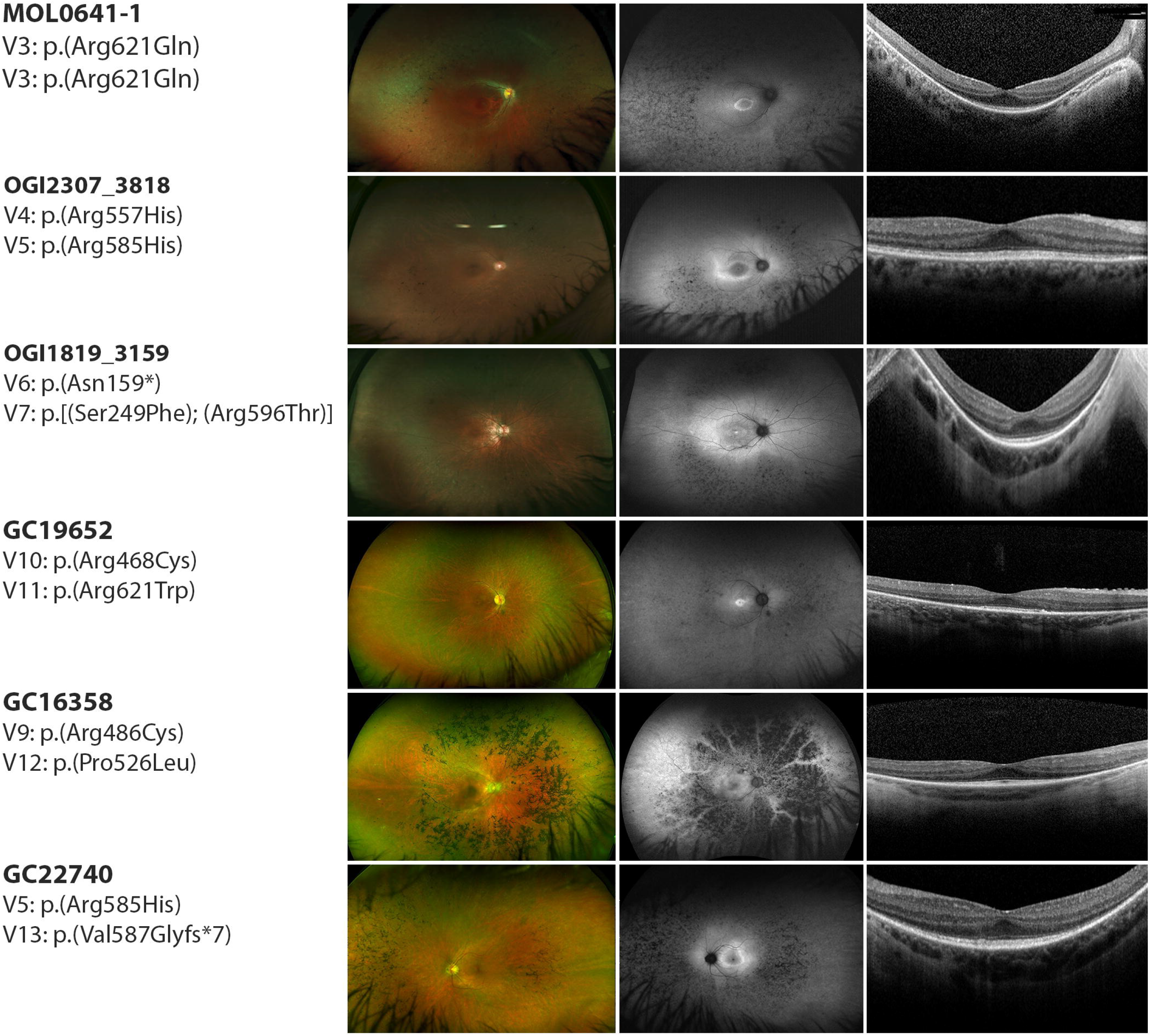
Clinical phenotypes of *INPP5E*-IRD patients. Images show fundus photos (left column), fundus autofluorescence (middle column), and OCTs (right column) for a representative subset of individuals.

**Table 1.**
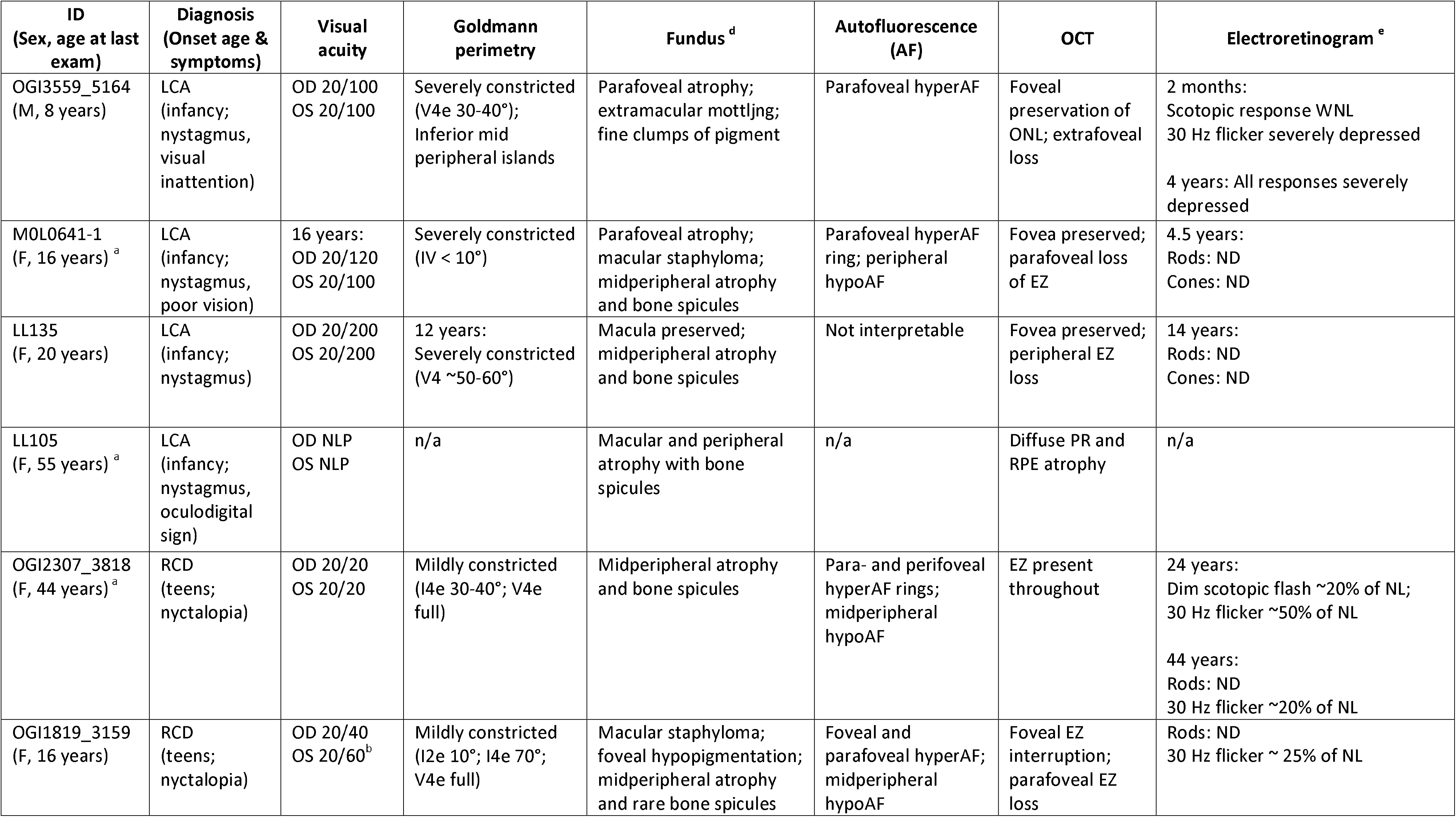
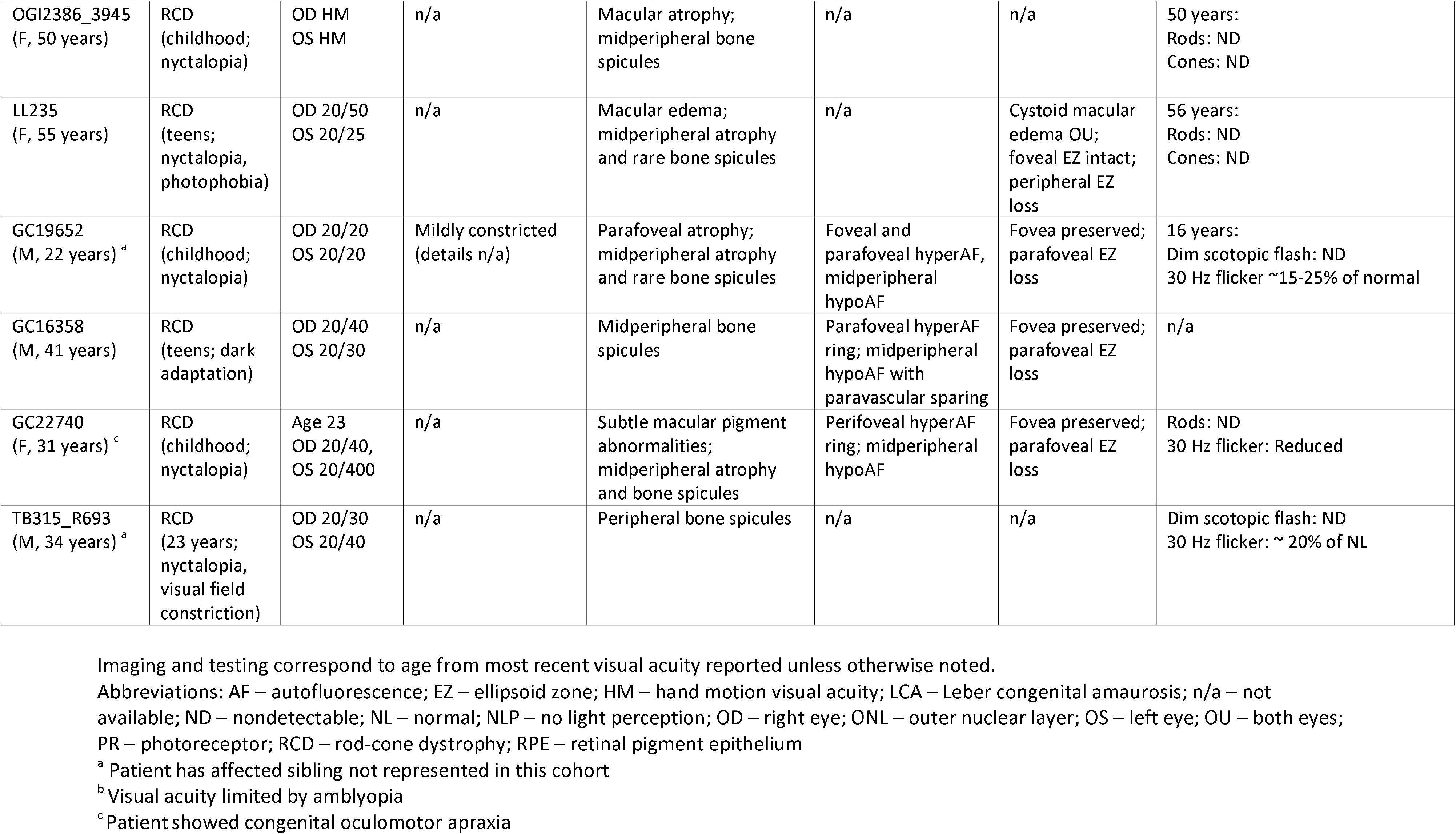
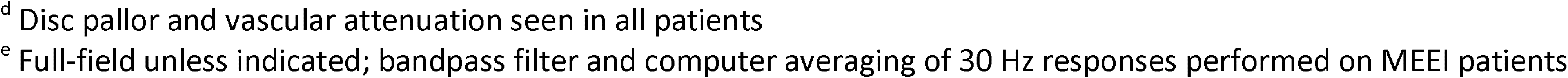

Individuals with RCD first experienced nyctalopia and impaired dark adaptation beginning typically in childhood and their teens. None of RCD subjects had nystagmus. Subjects had generally high visual acuities (Table 1). Goldmann perimetry showed mild constriction when available (n=3). Full-field ERGs were performed in seven of eight subjects with RCD. Scotopic responses were undetectable in all but one subject (OGI2307-3818 at age 24) whereas 30 Hz flicker (photopic) responses were present and relatively preserved in five patients (OGI2307-3818, OGI1819-3159, GC19652, GC22740, TB315_R693) (Table 1). Fundus examination and widefield fundus autofluorescence (FAF) imaging showed typical features of RCD in all individuals (Fig. 3, left and middle panel). Macular OCT imaging showed central ellipsoid zone (EZ) preservation in most patients, and in one (OGI2307-3818), the EZ was robust and identifiable through most of the scanned macula (Fig. 3). Bilateral cystoid macular edema was present in one individual (LL235). Most individuals in both groups for whom information about refraction was available were myopic.

Two subjects showed extra-ocular features: subject GC22740 presented with oculomotor apraxia and hypotonia at an early age which resolved, and the individual did not show any neurological or cognitive disability as an adult. Subject LL135 was found to have hypoplasia of the inferior cerebellar vermis on brain MRI at the age 18 during an investigation of headaches (Fig. S5). She had motor delay in infancy specifically delayed head control and sitting as well as “lack of strength,” frequent falls, and learning difficulties in childhood. Despite this history, other milestones including speech and walking were met at appropriate age. This subject completed secondary education at age 18. On a recent neurological examination performed at age 22, mild ataxia and tandem gait disequilibrium was noted. Renal ultrasound performed at age 15 showed no renal anomalies. The remaining ten subjects in this cohort did not have other extra-ocular features.

### Meta-analysis of all pathogenic *INPP5E* variants and their phenotypic correlation

Pathogenic variants in *INPP5E* can lead to a broad phenotypic spectrum ranging from severe ciliopathies to non-syndromic IRD.^8,11,12,31–34^ We hypothesized that differences in disease severity are caused by a more severe variant combination present in syndromic versus non-syndromic patients. Therefore, we gathered all known pathogenic variants (n=47) in syndromic and non-syndromic *INPP5E* cases and analyzed their potential effect on protein function (Fig. 2A, Fig.4, Table S5). First, we noticed that the difference in severity was not the result of a significantly higher frequency of loss-of-function (LoF) alleles in syndromic patients (9/68 alleles) compared to non-syndromic patients (3/34 alleles) (Table S5, Fig. 4A). Only three of 34 syndromic patients carried homozygous LoF allele and none of the IRD patients were homozygous for a LoF variant (Table S5, Fig. 4A). The remaining changes were missense variants, mostly within the inositol polyphosphate catalytic domain, with no apparent clustering based on the disease severity (Fig. 2A). Only five variants were shared between syndromic and non-syndromic cases (Table S5, Fig. 4A). Using protein modelling and variant deleteriousness prediction algorithms we determined the potential impact of each variant and each allelic combination on *INPP5E* function. Overall, we did not find significant differences in conservation or deleteriousness scores of variants and allele combinations between the syndromic and non-syndromic cases (Tables S4-S5, Fig 4B, Mann-Whitney test p-value>0.05). One extreme example is the p.Arg621Gln variant, which has been observed as a homozygote in two non-syndromic and one mildly syndromic IRD cases in this study and recently reported in one subject with JBTS with no retinal degeneration.^35^ These observations indicate that other genetic factors may play a role in the *INPP5E* disease manifestation.

**Fig. 4.**
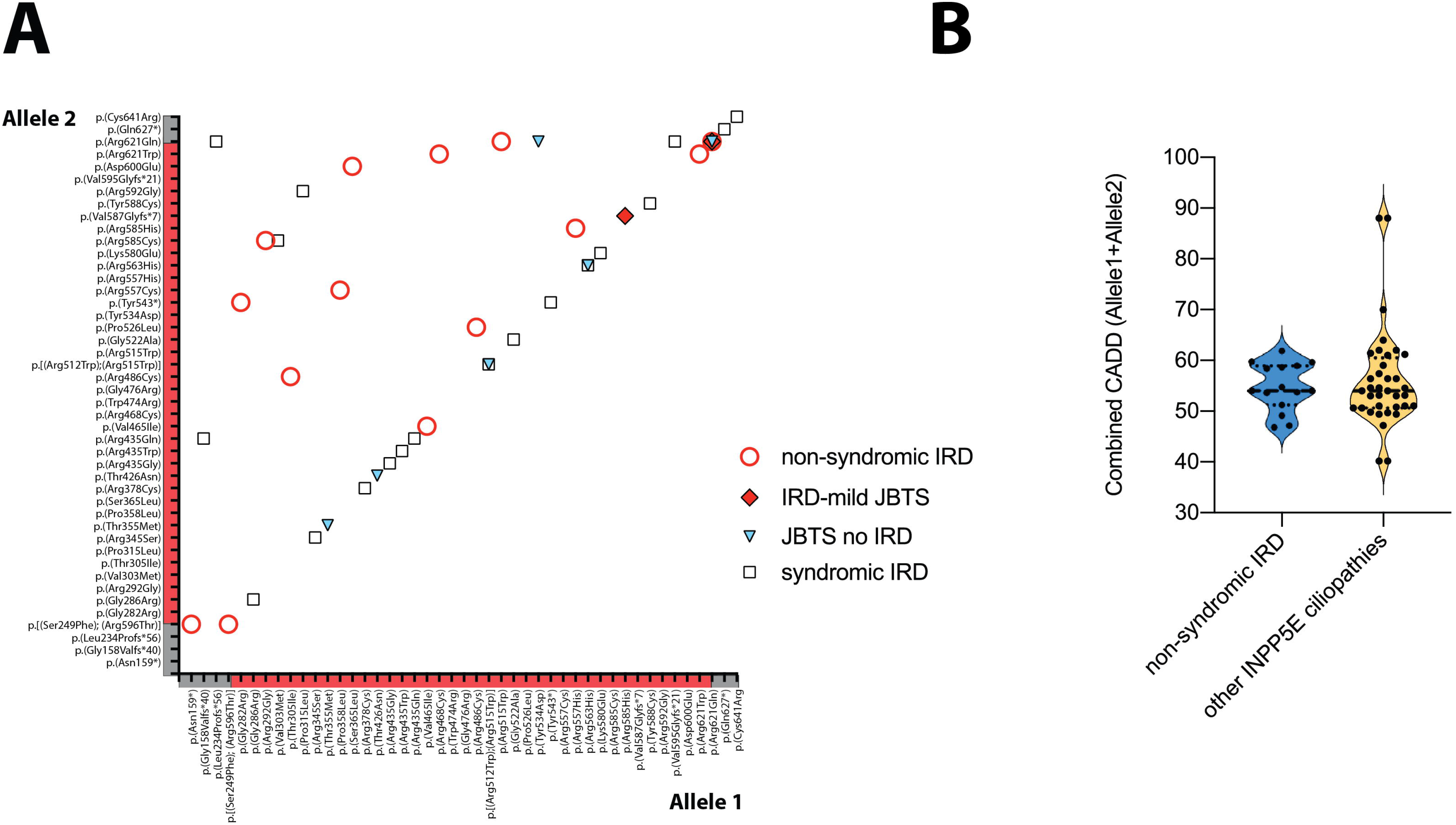
Meta-analysis of all pathogenic *INPP5E* variants and their phenotypic correlation. **(A)** Distribution of the *INPP5E* allele combinations in all reported INPP5E patients. Four different phenotypes of increasing severity were marked by circles (non-syndromic IRD), diamonds (IRD-mild JBTS), triangles (pure JTBS), and squares (syndromic IRD). **(B)** Violin plot of the cumulative CADD score for the *INPP5E* alleles in syndromic and non-syndromic IRD cases.

## DISCUSSION

Our study reveals a novel disease association of variants in *INPP5E* with non-syndromic retinal degeneration. We describe 12 mostly non-syndromic families, in which 18 affected members carried bi-allelic likely pathogenic variants in *INPP5E* resulting in phenotypes of LCA and RCD (or retinitis pigmentosa). Of the 14 alleles, 12 were novel and mainly resulting in missense changes of conserved amino acid residues in the phosphatase catalytic domain (Fig. 2A).

Pathogenic variants in *INPP5E* have been previously associated with systemic disorders, mainly JBTS, but were also seen in other ciliopathies.^8,11,12^ Sporadic IRD cases with pathogenic variants in *INPP5E* have also been reported in large mutational screening studies^31–34^, though clear association between *INPP5E* and non-syndromic IRD has never been established before. Ten of 12 probands reported here presented with vision problems with no other extra-ocular symptoms. Two cases, GC22740 and LL135, had mild ciliopathy features identified during childhood but as adults showed no neurological or cognitive disability. They were thus initially given a diagnosis of non-syndromic IRD. Hypoplasia of the inferior cerebellar vermis in LL135 was a secondary finding discovered by MRI performed to investigate the source of persistent headaches. Although this anatomical finding is less pronounced than the molar tooth sign in the classical JBTS, it is likely due to the *INPP5E* variants carried by this subject (Fig. S5). Since brain MRI was not performed on the remaining cases we cannot rule-out subclinical anatomical changes in these patients.

Two variants in our study, p.(Arg621Gln) and p.(Arg621Trp), affected the same residue. Both variants were predicted to disrupt a unique polar bond between Arginine 621 and a potential ligand. Homozygous p.(Arg621Gln) and p.(Arg621Trp) changes were found in five patients, three non-syndromic LCA patients (this study and ^31^), one mildly syndromic LCA case (LL135, this study), and one JBTS case without apparent retinal involvement.^35^ The p.(Arg621Gln) change has also been associated with non-syndromic IRD cases and with JBTS without retinal involvement in a compound heterozygous scenario.^29,33^ Unfortunately, at present the paucity of genotyped *INPP5E* patients makes it impossible to explain the phenotypic discrepancies in patients carrying the p.(Arg621Gln) variant. Nevertheless, the frequency at which Arginine 621 is mutated suggests that this amino acid constitutes a critical residue for the INPP5E function and together with the putative ligand binding by Arg621, warrants expansion of the catalytic domain of INPP5E to this position. Of the 19 known pathogenic alleles present in a homozygous state only three (p.[(Ser249Phe); (Arg596Thr)], p.(Val465Ile), p.(Arg621Trp)) resulted in a non-syndromic retinal degeneration, which may imply a hypomorphic or photoreceptor-specific impact of these variants on INPP5E function. These residues may also be important for photoreceptor-specific interactions with other ciliary proteins. Further functional studies will be needed to understand the impact of the identified *INPP5E* variants on the phosphatase activity or interactions with other proteins.

In order to understand the broad phenotypic spectrum of *INPP5E*-associated disease we used several deleteriousness prediction algorithms and protein modelling to uncover the impact of each variant on the protein function. We have not found significant differences between the syndromic and non-syndromic cases or between the LCA and RCD cases, analyzing the combined impact of both alleles in each patient. The lack of clear correlation of predicted variant impact on phenotype indicates that other genetic factors may play a role. Previous studies have shown that the INPP5E function in the cilium is dependent on other ciliary proteins, such as ARL3 and TULP3, and defects in those proteins lead to reduced or absent INPP5E localization to primary cilia.^36,37^ Moreover genetic modifiers in *cis* or *trans* to the primary disease variant(s) have been reported in many IRD studies where they influence disease penetrance, severity, and progression.^38^ For example, the *AHI1* allele p.(Arg830Trp) modifies the relative risk of retinal degeneration greater than seven fold within a nephronophthisis cohort.^39^ Similarly, resequencing of *TTC21B* gene in a large group of clinically diverse ciliopathies showed that variants in this gene account as severity modifiers in ^~^5% of ciliopathy patients.^40^ Although the number of genotyped samples with specific disease phenotypes are not large enough to support an unquestionable genotype-phenotype association, the rapid increase of high-throughput exome and genome sequencing in standard diagnostic protocols will help to validate some of these associations in the near future. In conclusion, we expanded the phenotypic spectrum of disorders due to pathogenic variants in *INPP5E* and demonstrated that these variants also account for a previously underdiagnosed form of non-syndromic IRD.

## Data Availability

The variants and their analyses described in the manuscript are listed in the manuscript figures and tables as well as the supplementary material.

## ACKNOWLEDGEMENTS

This work was supported by grants from the National Eye Institute [R01EY012910 (EAP), R01EY026904 (KMB/EAP) and P30EY014104 (MEEI core support)], the Jürg Tschopp MD-PhD Scholarship (VGP), the Swiss National Science Foundation [31003A_176097 (CR)], and the Foundation Fighting Blindness (EGI-GE-1218-0753-UCSD, KMB/EAP and BR-GE-0214-0639-TECH to TB, DS, and EB). Supported by grants from the National Institute for Health Research Biomedical Research Centre at Moorfields Eye Hospital NHS Foundation Trust and UCL Institute of Ophthalmology (RB, MM, ARW, GA), Moorfields Eye Charity (MM), and Retina UK (MM). The authors would like to thank the patients and their family members for their participation in this study and the Ocular Genomics Institute Genomics Core members for their experimental assistance. We specifically would like to thank Cassandra Amarello and Hilary Scott for their technical assistance. The authors would like to thank the Exome Aggregation Consortium, the Genome Aggregation Database (GnomAD) and the groups that provided exome variant data for comparison. A full list of contributing groups can be found at http://exac.broadinstitute.org/about and http://gnomad.broadinstitute.org/about.

## DISCLOSURE

The authors declare no conflicts of interest.

---

**Fig. S1 Variant validation and phasing in proband OGI1819_3159**

**Fig. S2 INPP5E protein alignment in 100 different species**. Evolutionarily conserved residues are indicated at the bottom of the alignment by using the symbols period (.), colon (:), and asterisk (*), consistent with increasing conservation. Clustal Omega predicted domain (residues 297-599 of human INPP5E) is indicated by a dark green bar and it overlaps with the Pfam-predicted phosphatase domain (residues 304-584). However, due to the significant conservation of region encompassing residues 273-296 and 600-621, we redefined the length of the INPP5E functional domain and indicated it by a light green bar. Residues mutated in the IRD probands presented in this study are highlighted in bold red.

**Fig. S3 Protein modelling of the known INPP5E catalytic domain**. One glycerol molecule (highlighted in green) is predicted to form polar bonds (cyan dashed lines) with residues His424, Asn479, Asp477, and His584. Three of the likely pathogenic variants identified in this study (highlighted in red) are located either within the catalytic pocket: p.(Arg557His) and p.(Arg585His) or in its proximity: p.(Thr305Ile).

**Fig. S4 Prediction of the structural changes caused by the p.(Thr305Ile) variant. (A)** The Thr305 is predicted to form polar bonds (cyan dashed lines) with residues Val587 and Gln339. **(B)** The p.(Thr305Ile) variant leads to the disruption of the polar bond with Gln339 but not of the ones with Val587.

**Fig. S5 MRI comparison between healthy subject, proband LL135 and patient with classical Joubert Syndrome**. MRI comparison between a healthy age and sex-matched subject **(A-C)**, proband LL135 **(D-F)**, and one patient with classical JS **(G-I)** demonstrated mild vermian hypoplasia (**D**, orange dashed line) on sagittal plane compared to JS patient who had severe vermian hypoplasia, also referred to as molar tooth sign (**G**, red dashed line). On coronal images, a dysplastic appearance of the superior vermian folia (**E**, orange arrow) was additionally seen, as in JS patient (**H**, red arrow). Compared to healthy subject who had short vertically oriented superior cerebellar peduncles (SCP, white arrow head), proband LL135 presented with thin elongated horizontally-oriented SCP (**F**, orange arrow head) and JS patient with very thick elongated horizontally-oriented SCP (**I**, red arrow head).

## Notes

### Competing Interest Statement

The authors have declared no competing interest.

### Author Declarations

The study was approved by the institutional review board of all participating institutions: Partners HealthCare System, the Boston Children Hospital Committee, Instituto de Oftalmologia, the Institutional Review Boards and ethics committees of Moorfields Eye Hospital, the institutional review board at Hadassah-Hebrew University Medical Center, the Ethics Comittee at Rambam Health Care Campus. The study adhered to the Declaration of Helsinki. Informed consent was obtained from all individuals on whom genetic testing and further molecular evaluations were performed.

